# Virtual Walking System with Mood Evaluation for Individuals with Severe Mobility Impairments: Development and Feasibility Study

**DOI:** 10.64898/2026.02.17.26346382

**Authors:** Yushen Dai, Yufei Lu, Yan Li, Mengqi Li, Ye Jia, Zihong Zhou, Chen Li

## Abstract

**Background:** Individuals with severe mobility impairments (SMI) often experience significant psychological distress and chronic pain. Virtual walking (VW) presents an innovative rehabilitation approach to improve mood and alleviate pain. This study aimed to develop a home-based VW system with integrated mood and symptom tracking and to report on its feasibility and usability in a user study with individuals with SMI.

**Methods:** A multidisciplinary, iterative framework guided the system’s development. Following initial contextual research and design iterations, a user study was conducted with 11 participants with SMI. A repeated measures pre-post design was employed. Feasibility and usability were primarily assessed through post-study qualitative interviews, analyzed via content analysis. Changes in mood and symptoms were measured immediately before and after each session. Momentary mood was captured using an in-virtual reality (in-VR) two-dimensional (2D) affect grid, while embedded single-item state ratings were used to track anxiety, depressed mood, and pain. Daily mood changes and symptom trajectories were analyzed using logistic regression and generalized estimating equations (GEE), respectively.

**Results:** Contextual research guided the system design towards enhancing accessibility, ergonomics, and therapeutic engagement. The final VW system featured three core modules: locomotion, multi-sensory feedback, and mood/symptom tracking. Qualitative analysis of the user study revealed high acceptance for the VW system, alongside challenges related to content variety and hardware ergonomics. Each intervention session was significantly associated with an immediate positive mood shift (odds ratio (OR) = 1.83), as measured by the affect grid. Furthermore, GEE models revealed a significant reduction in self-reported depression and anxiety symptoms over the intervention period (all *P* < 0.01).

**Conclusions:** This study confirms the feasibility and acceptability of the novel VW system for home-based use by individuals with SMI. The preliminary evidence suggests the system has high potential as a tool for improving mood and alleviating psychological distress. Future large-scale randomized controlled trials are warranted to establish its clinical efficacy.

**Trial registration number:** NCT07073144–07/17/2025.

## 1 Background

Severe mobility impairment (SMI), defined as an inability to walk or climb stairs independently, often necessitates the use of assistive devices and is associated with psychological distress, including anxiety and stress [1–3]. This psychological burden is often compounded by persistent pain, loss of independence, and reduced participation in daily activities [4, 5]. While pharmacological approaches (selective serotonin reuptake inhibitors, SSRIs) and conventional physical training are common management strategies for psychological issues, their utility can be limited by poor adherence, adverse side effects, and practical barriers, such as the risk of falls during physical activity [6–8]. There is a clear need for accessible, engaging, and safe approaches that can be deployed within an individual’s own living environment [9].

Virtual reality (VR) has emerged as a promising approach in rehabilitation, capable of stimulating complex, motivating environments that may enhance mood and support therapeutic goals [10]. For individuals with SMI, virtual walking (VW) systems that create the illusion of walking through an immersive virtual world offer a particularly compelling approach. The design of such systems involves critical modules, regarding hardware, locomotion methods, and the virtual environment itself, each presenting unique challenges for home-based deployment. A primary consideration is the hardware configuration used to deliver sensory feedback. While some systems use two-dimensional (2D) screens, head-mounted displays (HMDs) are increasingly favored for their ability to provide a wide field of view and a stronger sense of immersion [11]. However, many advanced VW systems incorporate peripherals that are poorly suited for home use; for example, haptic feedback is often delivered via specialized shoes or sensor-equipped floors that require tethering to external computers and technical expertise for setup [12, 13]. Similarly, hardware for motion-related feedback, such as treadmills or large omnidirectional stages, is often bulky, expensive, and impractical for a typical home environment [14, 15]. This has driven interest towards standalone HMDs that integrate visual, auditory, and controller-based haptic feedback in a single, portable package.

The method of locomotion is another core component that dictates system feasibility for users with SMI [11]. Passive observation of pre-recorded videos of a walking path with a 2D screen offers limited engagement and no user agency [16]. Another foot-tracking locomotion requires lower-limb motions, such as physical walking on a treadmill or foot-tracking systems, which are often not viable for this population due to physical limitations [17]. Consequently, controller-based locomotion is a common alternative [18]. Users control the analogue joysticks (e.g., HTC Vive and Oculus controllers) or press the trigger, which enables them to move in the virtual environment [19]. While joystick-based movement is functional, it can feel artificial and less immersive than gestures that mimic natural walking [19]. Arm-swing locomotion, where users move their arms to control the virtual avatar moving forward, offers a more intuitive and embodied experience [20]. However, existing arm-swing locomotion often requires large, simple virtual environments or depends on extra body-worn trackers, making it challenging for seated users or those with limited upper-limb function, thereby creating barriers to home-based deployment [15, 21].

Additionally, the design of the virtual environment and the integration of user feedback mechanisms are critical for engagement and therapeutic relevance [5]. Many existing VW systems employ simple, repetitive three-dimensional (3D) environments or offer only one or two scenes, which can lead to monotony [20]. Research indicates that exposure to virtual natural environments can relax participants’ mood, suggesting that providing a variety of rich, naturalistic scenes is a valuable design strategy [22]. Furthermore, few VW systems incorporate tools to track users’ mood states during a given session. The ability to capture brief, in-session data on momentary mood and symptoms like anxiety and pain could provide valuable, timely feedback for both users and clinicians; yet, this feature remains largely unexplored [23].

To address these gaps, we developed and evaluated a standalone, HMD-based VW system specifically optimized for home use by individuals with SMI. Our design combines an improved controller-based algorithm for the arm-swing locomotion technique designed for small and variable seated movements; synchronized multisensory feedback delivered through the HMD and its controllers; and an embedded evaluation module for brief in-session tracking of mood and symptoms within a variety of virtual natural environments. Guided by the Multidisciplinary Iterative Design of Exergames (MIDE) framework [24], we conducted a study to (1) define design requirements through contextual research, (2) develop and iterate on the VW system, and (3) evaluate its feasibility and user experience in a home-based setting.

## 2 Contextual Research (Phase 1)

### 2.1 Contextual Research

#### Accessibility for SMI

To inform the design of a VW system suitable for individuals with SMI, a scoping review of existing VW interventions was conducted. This review specifically aimed to identify: (1) prevalent locomotion methods, (2) upperlimb engagement considerations, and (3) strategies for minimizing cybersickness in this population [11].

#### Ergonomics and feasibility

A VR developer and a researcher in the mental health field scoped VR brands (Meta, Pico, and HTC) and identified mood monitoring and multisensory cues relevant for home use [22, 25].

#### Therapeutic needs

To understand the therapeutic needs of the VW system. First, we identified them through targeted searches of medical databases (Embase, PubMed) and grey literature (e.g., government health resources). The search strategy combined keywords related to the population (“severe mobility impairment” and “physical disability”), the intervention (“virtual walking”), and outcomes (“mood,” “well-being,” and “dosage”). Findings were synthesized to identify common recommendations for session duration, frequency, and psychological components (e.g., the need for mood relaxation). Then, these preliminary findings were discussed in a structured consultation with a senior clinician experienced in SMI rehabilitation to validate their clinical relevance and refine the design requirements. Identified needs, including delivery dosage and mood relaxation, were converted into design requirements through consensus within the research team and mapped to system features (Table 1). The adapted MIDE framework is depicted in Figure 1.

**Table 1.** Contextual Research: Key Design Inputs and Resulting Specifications.

| Domain | Contextual Research Finding | Design Goal | Design Specification (VW system & user study) |
| --- | --- | --- | --- |
| *Accessibility for SMI* |  |  |  |
| Locomotion | Higher immersion with arm swing | Develop an accessible locomotion method for SMI users | Arm-swing locomotion adapted for seated use |
| Upper limb | Limited upper-limb RoM | Identical to above | Locomotion is triggered via small wrist motions |
| Cybersickness | Need to minimize fatigue and nausea | Ensure high accessibility and minimize side effects | Stable frame rate; safety instructions |
| *Ergonomics & Feasibility* |  |  |  |
| Interface/Hardware | Home use favors standalone HMDs | Ensure feasibility for independent home use | Standalone HMDs (Quest 2/Pico 4) |
| Monitoring | Need for brief and feasible tracking | Integrate seamless and low-burden monitoring | Embedded single-item in-VR ratings for monitoring |
| Multisensory cues | Cues enhance presence | Enhance immersion and user experience | Scene-congruent audio & haptics |
| *Therapeutic Needs* |  |  |  |
| Dosage | Typical session: 11-15 mins | Establish effective therapeutic dose | 13-minute walking sessions |
| Frequency | Three to seven sessions/week | Identical to above | Daily sessions for ten days |
| Mood | Desire for mood relaxation | Promote mood relaxation | Natural environments |

*Note.* HMD = head-mounted display; RoM = range of motion; SMI = severe mobility impairments; VR = virtual reality; VW = virtual walking.

**Figure 1.**
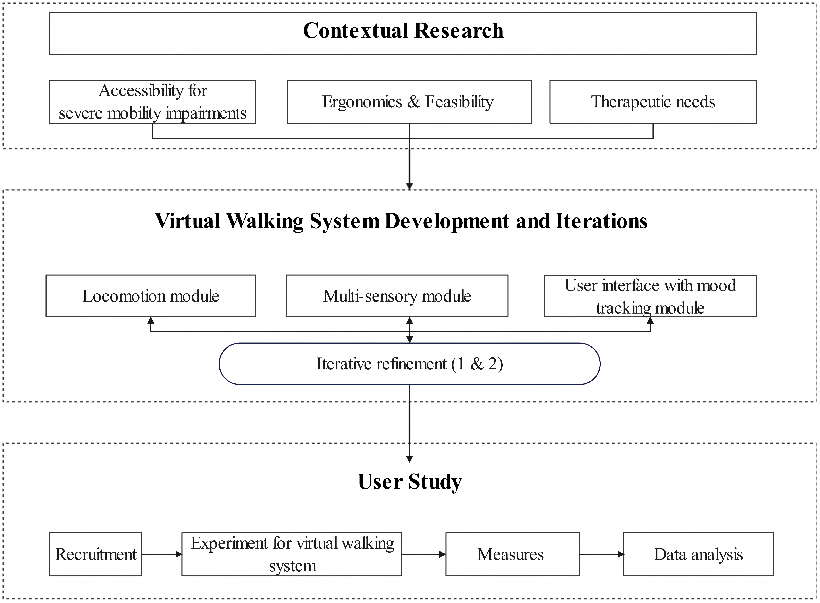
The adapted Multidisciplinary Iterative Design of Exergames framework

### 2.2 Results of Contextual Research

Regarding accessibility for individuals with SMI, the literature review indicated that arm swing locomotion mimics the arm movements of natural walking and can accommodate a range of upper-limb functions [11]; thus, we chose this locomotion method. We adopted built-in cybersickness mitigation strategies to minimize side effects and ensure accessibility [26].

Further ergonomic and feasibility analyses specified the use of Meta Quest 2 and Pico 4 headsets with hand controllers to provide multimodal sensory feed-back [27]. These multisensory cues included controller vibration during stepping and scene-congruent footstep sounds delivered through the HMD audio. For mood monitoring, the VW system was designed for momentary mood and single-item symptom evaluation (pain, anxiety, and depression) [11].

Informed by the therapeutic needs of individuals with SMI and results from the literature review, most protocols delivered sessions lasting 11–20 minutes, one to five times per week over 10–14 days [27–29]. As the optimal dosage is unknown, we specified a ten-day program with daily 13-minute sessions (ten minutes of VW and three minutes for preparation), approximating the mean dosage across the reviewed studies [11]. Virtual natural scenes and a first-person perspective avatar with visible lower limbs and synchronized leg movements were adopted to support mood relaxation and embodiment [30]. The detailed design goals and design specifications resulting from contextual research findings are summarized in Table 1.

## 3 Virtual Walking System Development and Iterations (Phase 2)

### 3.1 Virtual Walking Development

The VW system was developed in Unity3D (Unity Technologies, USA) by a VR developer. The system architecture comprised three modules derived from the findings of the contextual research (Table 1): (1) a locomotion module that generated walking from arm swing; (2) a multisensory module that delivered visual, auditory, and haptic cues to enhance presence; and (3) a user interface module with embedded mood and symptom tracking panels.

#### 3.1.1 Locomotion Module

##### Avatar integration

A generic humanoid avatar compatible with Unity’s Humanoid rig was used. A looping walking animation was parameterized by a normalized variable (0–1) that progressed from leg lift to foot placement. The avatar’s head and hands followed the positions and rotations of the user’s HMD and controllers, which were tracked by the headset’s inside-out tracking system. A first-person view with visible 3D hands and lower limbs was presented; leg movements were synchronized with user-driven locomotion to support a co-walking illusion.

##### Algorithm

Locomotion is driven by controller motion, building on prior armswing methods [20]. Figure 2 presents the algorithm for the locomotion module. The virtual origin, which serves as the main reference point linking the physical and virtual spaces, is placed at floor level directly below the user’s headset at the start of the session. To ensure stable yet flexible control for arm-swing locomotion, we implement a virtual three-dimensional “motion zone” in the virtual environment, defined as a bounding box with dimensions *x* (width), *y* (height), and *z* (depth), oriented to afford a wide horizontal range. This zone defines two operational states, each governed by the user’s hand controller positions. As illustrated in Figure 2, the user’s hand movement is defined by the angle *θ*, which represents the angle between the downward vertical axis originating from the point located at a horizontal distance *D* behind the virtual origin and a vertical height *H* above it and the line segment connecting this same point to the hand controllers. *θ*_L_ stands for the current angle of the left-hand controller, and *θ*_R_ stands for that of the right one. While both controllers remain within the “motion zone”, the system maintains a fixed spatial reference, keeping the initial reference angles (*θ*_L0_, *θ*_R0_) constant to provide a stable and predictable control for locomotion. The initial reference angles refer to the current angles captured at the moment when the user begins locomotion or when these angles are reset by the algorithm. A reset is triggered when either controller exits the zone boundary. Upon reset, the motion zone is algorithmically repositioned to the average spatial position of both the left and right controller positions. Concurrently, the reference angles (*θ*_L_, *θ*_R_) are updated based on the left- and right-hand controllers’ positions. This dual-state mechanism balances short-term control stability with long-term flexibility, allowing users to rely on consistent muscle memory for locomotion while freely adapting their posture and physical space.

**Figure 2.**
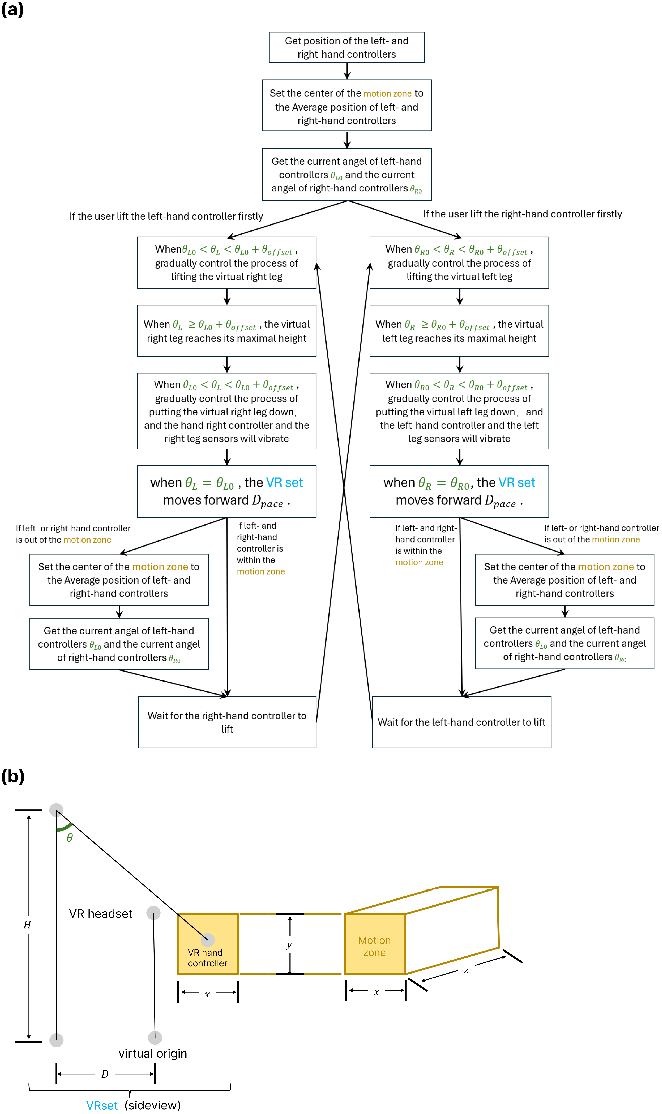
Locomotion algorithm (a) Flowchart of the algorithm and (b) Position diagram of virtual elements

For each hand, the walking animation is controlled by the angle between the controller and a reference point located behind and above the user (with a vertical offset *H* and a horizontal offset *D* ), defined relative to the virtual origin. As the angle increases, the corresponding leg is lifted until a threshold; the threshold is set as the initial reference angle within the motion zone plus an angle offset, *θ*_offset_. After exceeding the threshold, the leg lowers as the angle decreases, returning to the initial angle. Upon return, the avatar and VR set (as illustrated in Figure 2, where the VR set includes the virtual coordinates of the VR headset, the virtual origin, and the left- and right-hand controllers) advance discretely along a predefined path by a fixed step distance, *D*_pace_. Users alternate left- and right-hand motions to produce locomotion. Consistent with prior surveys on VR locomotion [20, 31], we refined our parameters during internal team testing to balance responsiveness with stability for seated users. The final parameters were set as follow: (*x* = 20 cm, *y* = 20 cm, *z* = 160 cm, *D* = 2 m, *D*_pace_ = 0.6125 m).

If a hand moves outside the motion zone, the zone’s center will be reset to the average position of both hands, and the initial angle is updated. This design aims to balance flexibility (variable hand placement, limited range of motion) with stability (reducing inadvertent triggering).

#### 3.1.2 Multisensory Module

Audio feedback included ambient environmental sounds and footstep effects to support immersion [32]. The ambient track switched among five soundscapes depending on the selected environment. Haptic feedback included controller vibrations and optional Bluetooth-connected foot vibration units (Tactosy for Feet, Pair).

#### 3.1.3 User Interface with Mood Tracking Module

We implemented a functional and notification user interface (UI) [33]. UI panels were presented as 3D planes. The functional UI allowed selection among ten walking scenes; hovering revealed additional information. Pre- and post-session mood rating panels included visual prompts (e.g., a blinking highlight on the avatar’s index finger) for trigger-based selection. The notification UI displayed program progress, with session progress markers shown at 50%, 75%, and 100% of each session.

### 3.2 Virtual Walking System Iterations

Two refinement cycles were conducted. The first iteration was driven by internal testing conducted by the multidisciplinary team to address: (1) basic controllability, (2) visual comfort, and (3) session timing. The second iteration incorporated formative feedback from two individuals with SMI, focusing on (1) locomotion methods and (2) tolerance of upper-limb limitations. Issues were logged, prioritized against contextual research requirements, and resolved through team consensus; implemented changes were documented in an iteration log.

An iteration log summarizing refinements across two development cycles is provided in Table 2. Key changes included improved first-person alignment, reduced visually induced motion (e.g., removal of scene shaking), automated session termination, and increased flexibility to initiate locomotion via smaller wrist motions. The recorded virtual walking demo is provided in Supplementary Material S1.

**Table 2.** Virtual Walking System Development: Iteration Log.

| Iteration | Input Source | Issue Identified | Change Implemented |
| --- | --- | --- | --- |
| First (team) | Ten-minute session | Imprecise arm swing | Implemented a direct mapping where right-arm swing triggers left-leg step, and vice-versa |
|  |  | Excessive animation | Removed shaking/effects |
|  |  | Unrealistic avatar | Refined proportions & alignment |
|  |  | No auto-timing | Added auto-termination |
| Second (SMI) | Ten-min session | Large arm range needed | Reduced required arm movement range by triggering locomotion via small wrist extension; anchored reference point behind the user's head to stabilize tracking |
|  |  | Difficult HMD fit | Assistance for IPD/headband |

*Note.* HMD = head-mounted display; IPD = interpupillary distance; SMI = severe mobility impairments.

## 4 User Study (Phase 3)

### 4.1 Experiment Design and Setting

The user study followed a ten-day repeated pre–post design. Users used the VW system in their homes. However, an initial onboarding session for each participant was conducted in a university meeting room, where a research assistant provided training, reviewed safety procedures, and supervised the user’s first VW session. During this session, the user completed their first 13-minute VW session before taking the equipment home for subsequent self-administered sessions. The study was approved by the Research Ethics Committee of the Hong Kong Polytechnic University, and all users provided written informed consent.

### 4.2 Users

Users were recruited through convenience sampling in February 2025. A formal sample size calculation was not conducted due to the nature of this development study.

A target sample of 10–12 participants was deemed sufficient to achieve data saturation for qualitative feedback and identify major usability issues of the VW system [34]. SMI was operationalized as the inability to walk four meters in ten seconds, self-/proxy-/record-reported inability to walk across a small room, or dependence on a walking aid for *>* 6 months [2]. Inclusion criteria were: (1) age 18–65 years; (2) ≥ 6 months post-injury or disease onset and clinically stable; and (3) Cantonese-speaking. Exclusion criteria were: (1) current severe psychiatric disorder (e.g., schizophrenia, major depression) and (2) severe cognitive impairment (Abbreviated Mental Test score *<* 6 [35]).

### 4.3 Experiment Protocol

Users received one daily VW session lasting 13 minutes using a Pico 4 or Quest 2. Users could select one virtual nature scene per session. The duration of each session was pre-designed in the system. After each session, the system would exit automatically. Users were provided with a paper pamphlet to record session duration and any side effects after each session. The log application on the device records timestamps and session durations. A research assistant provided training on the VR system usage to users and verified safety procedures before they used it. If there were missed days, the research assistant would provide an additional two days for users to complete the entire experiment. Technical questions were addressed via WhatsApp.

### 4.4 Measures

#### User experience evaluation

Semi-structured interviews were conducted to explore system performance and home usability (see Supplementary material S2 for the full interview guide). One-to-one interviews were conducted face-to-face or online. This was designed to generate practice-relevant insights for further refinement of the system, rather than to provide definitive feasibility estimates for a trial.

#### Embedded mood and symptom tracking

Mood and symptoms were measured immediately before and after each session. Momentary mood was captured using a two-dimensional affect grid based on Russell’s circumplex model [36]. The grid represented valence (very unpleasant to very pleasant) and arousal (very calm to very activated), with discrete emotion labels (e.g., happy, calm, relaxed, anxious, sad) placed according to their valence–arousal coordinates. Participants selected one emotion label at the pre-session and post-session.

Embedded single-item state ratings were used to track anxiety, depressed mood, and pain. State anxiety and depressed mood were rated with the questions: “Right now, how anxious do you feel?” and “Right now, how depressed do you feel?”, scored from 0 (“not at all”) to 10 (“extremely”) [37]. Pain was rated using the Numeric Pain Rating Scale from 0 (“no pain”) to 10 (“worst possible pain”) [38]. These brief state ratings were designed for in-session monitoring and are not intended to replace validated clinical questionnaires.

### 4.5 Data Analysis

Quantitative analyses were conducted in R (v4.4.2) and IBM SPSS 26. The characteristics of the users were summarized as mean (SD) or *n* (%). For momentary mood, the emotion selections on the mood grid were dichotomized into positive (e.g., happy, calm, relaxed) vs. non-positive (neutral or negative) categories. For each intervention day, we derived a binary indicator of mood change (positive/maintained positive vs. non-positive change) from pre- to post-intervention. We then fitted mixed-effects logistic regression models with day (1–10) as a fixed effect. Single-item ratings were analyzed using GEE with an identity link and robust standard errors, including phase, day, and the phase × day. The level of significance was set at two-sided *α* = 0.05.

Qualitative data were managed in NVivo 20. A deductive content analysis was conducted, guided by predefined domains relating to usability and acceptability. Transcripts were divided into condensed meaning units and coded independently by two researchers. The researchers then met to discuss coding discrepancies and reach a consensus on shared coding. Codes were subsequently grouped into sub-categories and themes.

### 4.6 Results of User Study

Eleven users were enrolled (mean age 58.92 years, SD 8.20). Six users (54.55%) were female. Users reported basic to moderate computer experience and little to no VR exposure. All users had neurological conditions (e.g., poliomyelitis, spinal cord injury). The multidisciplinary team (mean age 31.75 years, SD 5.62) participated in the system design (see Table 3).

**Table 3.** Users’ Characteristics at Baseline by Group Assignment, *n* (%)

| Characteristic | SMI Users ($n = 11$) | Clinical Experts/VR Developer ($n = 4$) |
| --- | --- | --- |
| Age (years), mean (SD) | 57.90 (9.41) | 31.75 (5.62) |
| Sex |  |  |
| Male | 5 (45.45) | 2 (50.00) |
| Female | 6 (54.55) | 2 (50.00) |
| Computer experience | Basic to moderate | Moderate to expert |
| VR knowledge | None to basic | Basic to expert |
| Neurological disorder<sup>†</sup> | 11 (100.00) | — |

*Note.*<sup>†</sup>Neurological disorders include poliomyelitis, spinal cord injury, stroke, and cerebellar atrophy; (—) = not applicable; SD = standard deviation; SMI = severe mobility impairment; VR = virtual reality.

#### 4.6.1 User Experiences

All users completed the ten-day program. No serious adverse events occurred. Two themes were identified: (1) Generally well accepted, and (2) Ease of use and usability challenges.

Theme 1: Generally well accepted. Assessment of system feasibility revealed a high level of acceptability; users reported strong engagement stemming from the novelty of simulated locomotion (e.g., “rediscover the feeling of walking again,” P1). However, constraints regarding long-term engagement were identified, specifically centered on environmental content diversity, as limited scene realism led to perceived monotony (P3–P5; e.g., “The scene remained almost unchanged,” P3). Additionally, tolerability barriers included upper limbs and neck fatigue, defining the practical limits for the VW system (P4–P6; e.g., “I find my hands get very tired,” P4).

Theme 2: Ease of use and usability challenges. The system’s interaction design was generally characterized by a low barrier to entry, although initial navigational orientation required a brief learning phase (P2, P5–P6; e.g., “When using it for the first time, I may feel unaccustomed to the spatial awareness, but gradually you can realize this system isn’t complicated,” P6). HMD weight and cross-modal synchronicity. Specifically, a perceived mismatch between physical upper-limb swing and virtual locomotion feedback (e.g., “my arms and the walking on the screen sometimes felt out of sync,” P5) was identified as a key area for technical optimization.

#### 4.6.2 Embedded Mood and Symptom Tracking Results

##### Momentary mood

Across all sessions, most pre- and post-session selections fell within the high-valence quadrants of the affect grid (high-valence/high-arousal and high-valence/low-arousal). Transitions were more frequently observed within these high-valence quadrants than from high- to low-valence quadrants (Figure 3). In mixed-effects logistic regression models comparing positive versus non-positive mood change from pre- to post-intervention, the day showed a statistically significant positive mood change (*P* < 0.05). Each intervention day was associated with higher odds of experiencing a positive mood or a maintained-positive mood change (OR = 1.83, 95% CI = 1.10 − 3.03) (Table 4).

**Table 4.** Logistic Regression Results of Momentary Mood Evaluation.

|  | Estimate (SE) | Z | P | OR (95% CI) |
| --- | --- | --- | --- | --- |
| Intercept | 4.81(3.14) | 1.53 | 0.126 | 78.697 (0.261-57845.016) |
| Day | 0.60 (0.26) | 2.34 | 0.019* | 1.828 (1.103-3.032) |

*Note.* CI = confidence interval; GEE = generalized estimated equation; OR = odds ratio; SE = standard error; \* $P < 0.05$ .

**Figure 3.**
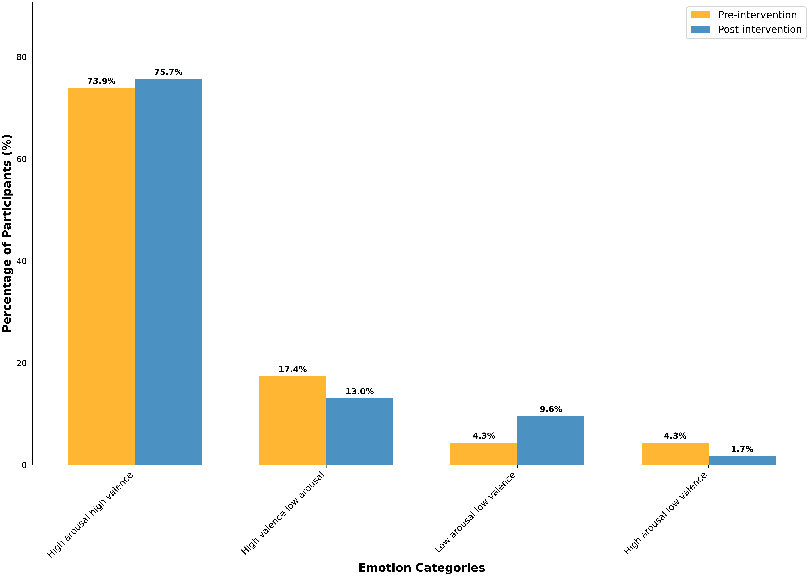
Pre- to post-experiment comparison between emotion quadrants among users

##### Single-item evaluations

GEE models revealed significant main effects of day in depression (*P* < 0.001), anxiety (*P* < 0.01), and pain ratings (*P* < 0.001) over the ten-day program. Statistically significant phase × day differences were observed in depression and anxiety ratings (*P* < 0.01) (see Table 5).

**Table 5.**
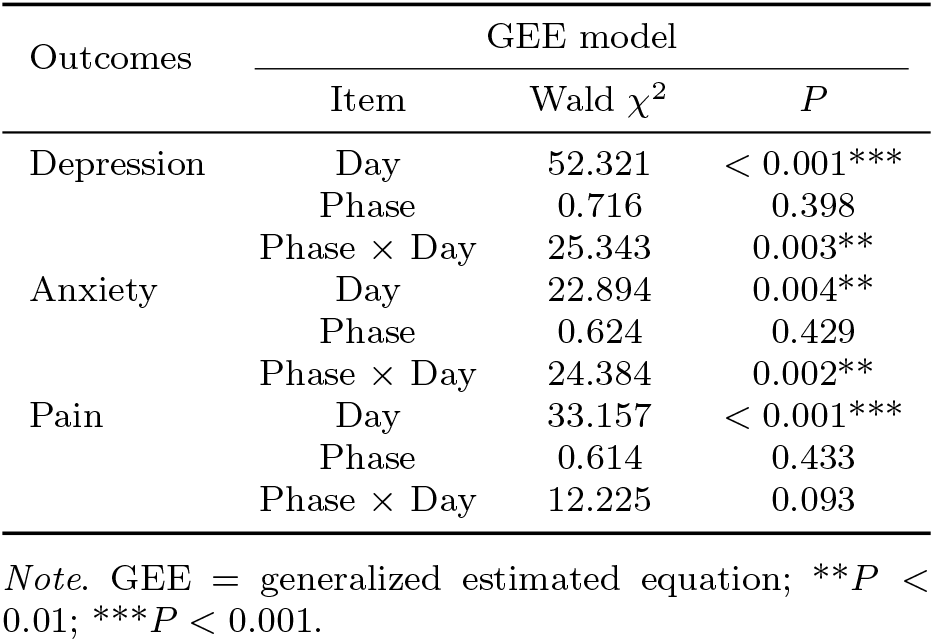
GEE Results of Single-item Questionnaire (Depression, Anxiety, and Pain)

| Outcomes | GEE model |  |  |
| --- | --- | --- | --- |
| | Item | Wald $\chi^2$ | $P$ |
| Depression | Day | 52.321 | < 0.001*** |
|  | Phase | 0.716 | 0.398 |
| | Phase × Day | 25.343 | 0.003** |
| Anxiety | Day | 22.894 | 0.004** |
|  | Phase | 0.624 | 0.429 |
| | Phase × Day | 24.384 | 0.002** |
| Pain | Day | 33.157 | < 0.001*** |
|  | Phase | 0.614 | 0.433 |
| | Phase × Day | 12.225 | 0.093 |

*Note.* GEE = generalized estimated equation; \*\* $P$ < 0.01; \*\*\* $P$ < 0.001.

## 5 Discussion

In this development-stage study, we used an adapted MIDE framework to design and evaluate the VW system for individuals with SMI. We translated design requirements from contextual research into a seated arm-swing locomotion algorithm, synchronized multisensory feedback, and embedded mood and symptom evaluation, and examined how this system functioned in real home use. High session completion rates and generally positive interview feedback indicate that the protocol and hardware-software configuration were acceptable for most users. However, qualitative data highlighted technical and design issues that may limit longer-term use.

Users frequently described the system as novel at the beginning of the program, despite their limited physical mobility. This aligns with prior HMD-based VW studies in non-clinical samples that reported strong initial engagement during nature-based walking [15, 39]. At the same time, the data indicate that environmental and hardware factors tightly influence feasibility at home. Some users reported intermittent, “disjointed” walking when hand tracking or controller motion became unstable. In a locomotion-based system, tracking instability directly reduces the coherence of the walking illusion and can disrupt presence [40]. Inside-out HMDs are known to be sensitive to lighting conditions, reflective surfaces, and occlusion [41], and these factors are harder to control in unstructured home environments than in lab settings. From a system design perspective, this stresses the need for in-app setup guidance (e.g., detection and feedback about poor lighting and unstable seating position) and the robustness of locomotion mapping to noisy motion inputs.

Users generally found the system easy to use once set up, but several usability barriers could limit sustained use: headset weight and fit, controller-based navigation demands, and the need for initial adjustment. These findings highlight that, for users with SMI, ease of use also includes physical handling, setup difficulties, and fit to the user’s function. While HMDs provide stronger immersion and support for home use [29, 42], these benefits come at the cost of added weight, pressure points, and sensitivity to individual head and neck limitations. Future iterations should consider hardware-software optimization, including reducing users’ manipulation demands (e.g., fewer controller buttons); minimizing repetitive arm lifting, and streamlining the initial adjustment pipeline (e.g., guided fitting, automated comfort check).

The locomotion module was designed to detect small forward swings of the controllers and map them to discrete steps, enabling seated users with limited upperlimb function to experience walking with relatively low effort. Unlike many existing arm-swing approaches that rely on simple direction thresholds or large front–back excursions relative to the head [28], our algorithm implements a “motion zone”, which is a virtual 3D box (the size was refined during internal team testing to 20 cm tall × 20 cm deep × 160 cm wide) in the virtual environment and balances stability and adaptability. The position of the “motion zone” remains fixed while both controllers are within its volume, ensuring a consistent control reference. A reset is triggered when a controller exits the zone, at which point the entire zone is instantly repositioned to the midpoint between the two controllers. This approach maintains predictable control during steady use while accommodating larger shifts in the user’s hand position. Qualitative feedback suggests that this approach was workable for most users, but it also revealed boundary conditions. Some users reported episodes where on-screen locomotion felt out of synchronization, even after we adjusted the size of the “motion zone”.

In real homes, factors such as variable seating posture, trunk compensation, and inconsistent arm range appear to challenge any fixed-parameter locomotion mapping [40]. Our findings highlight the need for algorithms that can adapt to smaller, noisier motion patterns over time. For example, future versions could estimate each user’s comfortable movement space during a short calibration phase and continuously adapt sensitivity and motion-zone parameters as usage continues [43]. In addition, combining controller data with head or torso motion and applying basic plausibility checks may help stabilize gait perception when tracking is degraded [44].

The VW system was safe to use as no serious adverse events occurred; only eye and arm fatigue were reported, aligning with prior VR studies and underscoring the importance of proactive comfort management in home-based protocols [28, 42, 45]. For home delivery, brief screening (e.g., Simulator Sickness Questionnaire (SSQ) via a short demo) may help identify users prone to cybersickness [25]. Practical strategies (comfort fit, optional breaks, and movement guidance tailored to limited mobility) could help reduce fatigue and dizziness [25]. Additionally, comfort management needs to be integrated into the usage model, particularly for home use.

Our use of embedded, single-item ratings prioritized feasibility for in-session tracking over psychometric depth [46]. Thus, these quantitative data should be seen as exploratory signals of user response, not as evidence of clinical effect. While the data suggest a trend towards positive mood changes, the clinical significance is constrained by a significant floor effect: many users reported minimal distress at baseline, limiting the potential for improvement. This highlights a key challenge for universal mood tracking in heterogeneous populations. Similarly, the lack of between-group differences in the general pain rating was unsurprising, as a single numeric scale cannot capture the distinct mechanisms (e.g., neuropathic vs. nociceptive) underlying SMI [47]. Future research should incorporate validated multi-item questionnaires and mechanism-specific pain assessments to draw meaningful clinical conclusions.

### Strengths and limitations

From a neuroengineering perspective, this work contributes a seated arm-swing locomotion algorithm explicitly tuned for small, variable upper-limb movements for home use in people with SMI. Additionally, the system integrated in-system symptom evaluation to track brief changes in depression, anxiety, and pain. However, this study has some limitations. First, the small, heterogeneous sample and single-group design mean that the findings cannot be interpreted as indicative of clinical efficacy. Second, the use of single-item measures and a simple affect grid favored feasibility over depth, limiting inferences about specific mechanisms. Finally, the locomotion and interaction modules require further optimization for long-term use in diverse home environments.

## 6 Conclusion

In conclusion, this study provides insight into the development and feasibility of a VW system for individuals with SMI using an adapted MIDE framework. The system appeared feasible and generally usable as a home-based option for managing mood and pain in this population. However, several aspects require further refinement and evaluation, including more precise tracking of the walking animation and conducting a randomized controlled trial to test clinical effectiveness rigorously.

## Abbreviations

CI: Confidence Interval
GEE: Generalised Estimating Equation
HMD: Head-Mounted Display
IPD: Interpupillary Distance
MIDE: Multidisciplinary Iterative Design of Exergames
OR: Odds Ratio
RoM: Range of motion
SD: Standard Deviation
SE: Standard error
SMI: Severe Mobility Impairment
SSQ: Simulator Sickness Questionnaire
SSRI: Selective Serotonin Reuptake Inhibitor
UI: User Interface
VR: Virtual Reality
VW: virtual walking

## Acknowledgements

We showed our gratitude to the users for their support and collaboration in this study.

## Declarations

### Ethics approval and consent to participate

Ethics approval was obtained from the Institutional Review Board of the Hong Kong Polytechnic University (No. HSEARS 20240930009), and all users signed the consent form.

### Consent for publication

Not applicable.

### Data availability

Data will be kept confidential only upon inquiry by the corresponding author.

### Materials availability

Not applicable.

### Code availability

Data will be kept confidential only upon inquiry by the corresponding author.

### Author contribution

YD: Research Design, Conceptualization, Writing-original draft, Writing-review & editing. YL: Project administration, Project development, Data collection, Writing-review & editing. YL: Project administration, Research design, Writing-review & editing. ML: Writing-review & editing. YJ: Project administration & Research design. ZZ: Writing-review & editing. CL: Project administration & Research design

