## Supplementary materials 2 for "Virtual Walking System with Mood Evaluation for Individuals with Severe Mobility Impairments: Development and Feasibility Study"

**Acceptability**

1. Which parts of the system are you satisfied with?

**Probing**:

(1) Setting up the equipment (e.g., HMD, Controller)

(2) The VR system (e.g., HMD, user interface, scenery, avatar, audio, locomotion method, haptics)

(3) Operational process (e.g., starting, pausing, finishing sessions)

2. Which parts of the system are you dissatisfied with?

**Probing:** Same as above

3. What aspects motivated you to continue using the system?

4. What aspects discouraged you from using it?

5. Would you like to continue using the system in the future, or recommend it to others? Why or why not?

6. Did you experience any discomfort while using the virtual walking system?

**Usability in daily use**

1. How **easy or difficult** was it to use the system at home for the first time?

2. Was the virtual walking process convenient or complicated? For example, how quickly could you start or set up each session? What was the shortest time it took you to begin walking?

3. Did you encounter any problems or make any mistakes while using the system at home? (e.g., getting stuck, misoperation)

4. What factors helped or made it difficult for you to use the system at home? (e.g., space, family, noise, etc.)
